# Effects of combining constraint-induced movement therapy and action-observation training on upper limb kinematics in children with unilateral cerebral palsy: A randomized controlled trial

**DOI:** 10.1101/19009779

**Authors:** Cristina Simon-Martinez, Lisa Mailleux, Ellen Jaspers, Els Ortibus, Kaat Desloovere, Katrijn Klingels, Hilde Feys

## Abstract

Modified constraint-induced movement therapy (mCIMT) improves upper limb (UL) motor execution in unilateral cerebral palsy (uCP). As these children also show motor planning deficits, action-observation training (AOT) might be of additional value. Here, we investigated the combined value of AOT to mCIMT on UL kinematics in children with uCP. Thirty-six children with uCP completed an UL kinematic evaluation after participating in a 9-day mCIMT camp wearing a splint for 6 hours/day. The experimental group (mCIMT+AOT, n=20) received 15 hours of AOT, i.e. video-observation and execution of unimanual tasks. The control group (mCIMT+placebo, n=16) watched biological-motion free videos and executed the same tasks. We examined changes in motor control (movement duration, peak velocity, time-to-peak velocity, and trajectory straightness) and movement patterns (using Statistical Parametric Mapping) during the execution of three unimanual, relevant tasks before the intervention, after and at 6 months follow-up. Adding AOT to mCIMT mainly affected movement duration during reaching, whereas little benefit is seen on UL movement patterns. mCIMT, with or without AOT, improved peak velocity and trajectory straightness, and proximal movement patterns. These results highlight the importance of including kinematics in an UL evaluation to capture changes in motor control and movement patterns of the proximal joints.

## 1. Introduction

The upper limb (UL) motor and sensory deficits in children with unilateral Cerebral Palsy (uCP) hinder their performance of daily life activities ^1^. Effective rehabilitation approaches include bimanual training ^2^, modified constraint-induced movement therapy ^3^, or goal-directed training ^4^. One of the most popular treatment modalities amongst clinicians and researchers is constraint-induced movement therapy (CIMT) and its modified versions (mCIMT) ^5^, due to its proven high effectiveness and its ability to improve both unimanual and bimanual function ^6–9^. mCIMT consists of constraining the less impaired hand while targeting intensive unimanual task-related practice with the more impaired UL ^10^.

Apart from motor execution problems, children with uCP may also have difficulties in motor representation and motor planning ^11,12^. These deficits can be targeted with Action-Observation Training (AOT), a novel treatment approach aimed at activating the mirror neuron system. Mirror neurons are a particular class of visuomotor neurons that discharge both when we execute a particular motor action, and when we observe another individual executing the same action ^13^. Through observation, we learn how to imitate the movements required to perform a specific action, which can be translated into skill learning. A few studies have already shown the efficacy of AOT in improving UL sensorimotor deficits in children with uCP ^14–18^. However, the added value of AOT to a well-established rehabilitation approach, such as mCIMT, has not yet been investigated.

Changes in UL sensorimotor function following mCIMT or AOT have been typically evaluated with clinical scales that rely on an ordinal-based (subjective) scoring system. Three-dimensional motion analysis (3DMA) provides a quantitative measurement of UL movement patterns of proximal and distal joints ^19–21^, highlighting its added value compared to clinical scales ^22^. Its utility and sensitivity to identify improvements at joint level after UL surgery and botulinum toxin injections has been shown ^23,24^. These improvements were most obvious in the proximal joints (shoulder, scapula or trunk) for which clinical scales are less sensitive ^23^. The analysis of waveforms derived from 3DMA data using Statistical Parametric Mapping (SPM1d) has allowed us to accurately map UL deficits in children with uCP ^25^. Treatment-induced changes on UL movement pathology require a comprehensive analysis over the entire waveform of the joint angle, and the use of SPM1d on 3DMA data will potentially enable us to capture changes in movement patterns after an intensive training remains unknown.

Although both mCIMT and AOT have individually been shown to be efficient in improving UL function in children with uCP, it remains unknown whether their combination would be more effective than mCIMT alone. Furthermore, the comprehensive evaluation of UL movement patterns obtained with 3DMA could be highly beneficial to capture training-induced changes, especially at the joint level. Therefore, the aim of this study was to examine the added value of AOT to mCIMT on improving UL movement patterns, as measured with UL-3DMA.

## 2. Results

### 2.1. Participants

Forty-four children with uCP participated in this study (mean age (SD) 9y6m (1y10m); 27 boys; 9 MACS I, 15 MACS II, 20 MACS III), and were randomized into the mCIMT+AOT (n=22) and the mCIMT+placebo (n=22). All children completed the intervention. In the mCIMT+AOT group, data of two children were lost immediately after the intervention. In the mCIMT+placebo group, data of four children were lost immediately after the intervention and data of two were lost to follow-up, resulting in a total of 36 children of whom n=20 received mCIMT+AOT and n=16 mCIMT+placebo (Figure 1). Baseline characteristics of the participants are reported in Table 1. All children who received AOT showed a good compliance to the video observation, based on the number of correct answers to the video-related questions (median=42/45 correctly answered, interquartile range=5, range 30-45).

**Table 1.**
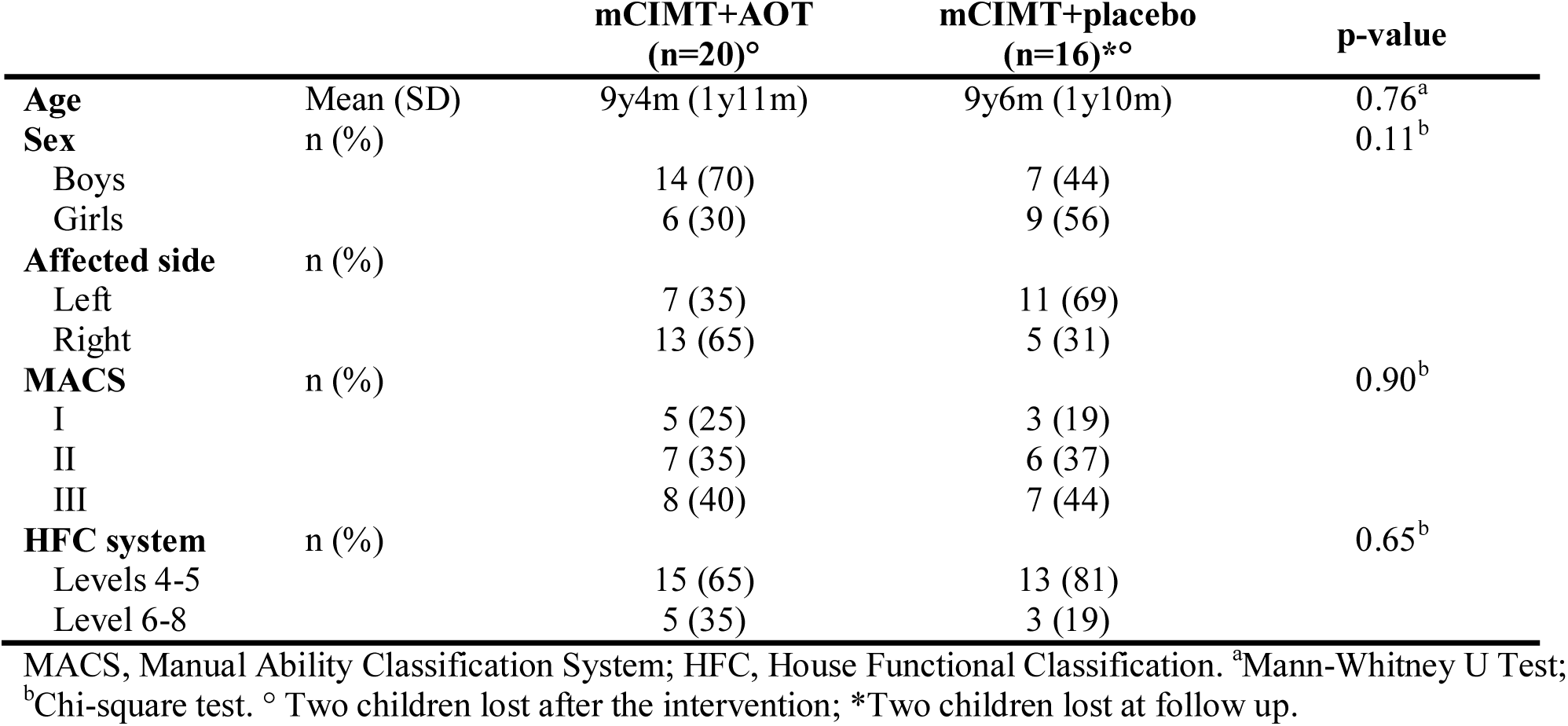
Descriptive characteristics of the participants per group and statistical comparison.

**Figure 1.**
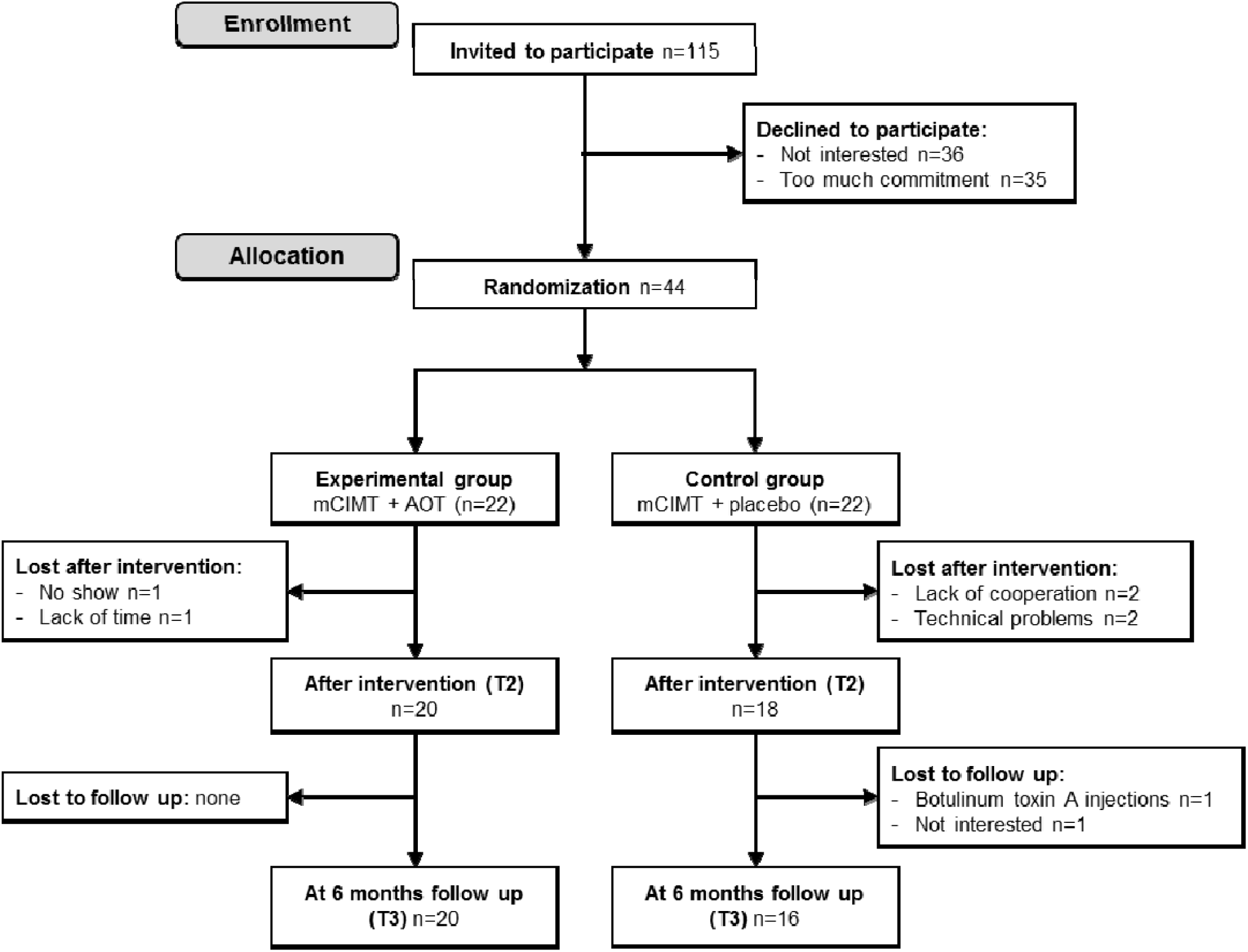
CONSORT flowchart with number of participants and reasons for missing data in each group, at each time-point.

### 2.2. Treatment efficacy

The effects of this RCT will be first reported for between-group differences to evaluate the added value of AOT to mCIMT. Next, the within-group effects over time will be described for the total group, to evaluate the effects of mCIMT with or without AOT.

#### Between-group analysis

Table 2 summarizes the improvement in motor control for each group (mCIMT+AOT vs. mCIMT+placebo) at every time point (T1, T2, and T3), revealed by the spatiotemporal parameters. We found no between-group differences at baseline (T1) for any spatiotemporal parameter (all p>0.05). We found a significant time*group interaction for movement duration during RU (F=3.37, p=0.04, partial η^2^=0.17) in favour of the mCIMT+AOT group (F=9.83, p=0.001). In this group, post-hoc analysis showed significant improvement at follow-up (T1-T3, p=0.001, d=0.64) (Figure 2). The mCIMT+placebo group showed no significant improvements in movement duration (p>0.05). No significant interactions were found for the other spatiotemporal parameters or the other tasks (p>0.05).

**Table 2.**
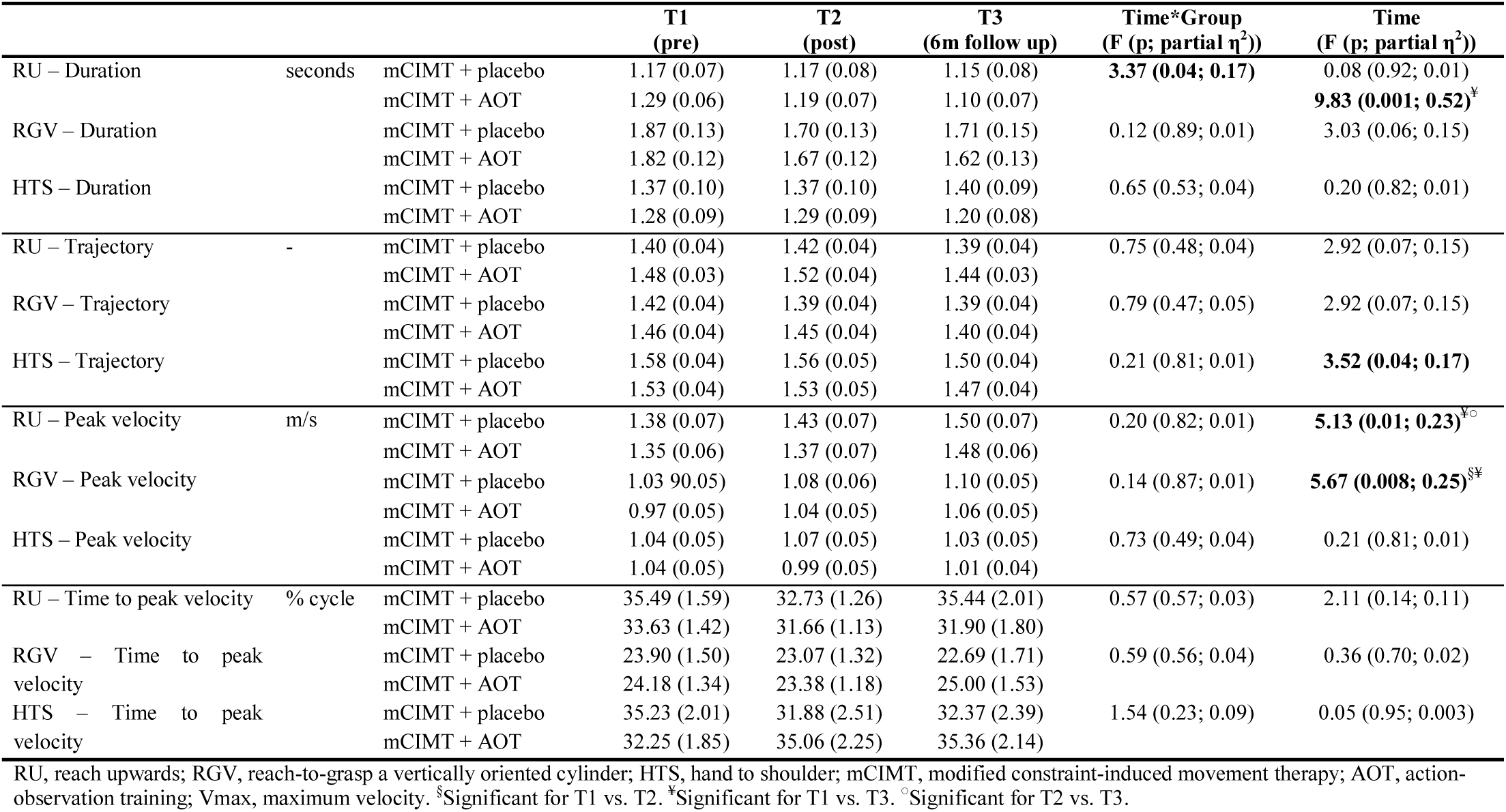
Estimated marginal means (standard error) of motor control parameters at each time-point, and statistical comparison (F (p-values; effect size)).

**Figure 2.**
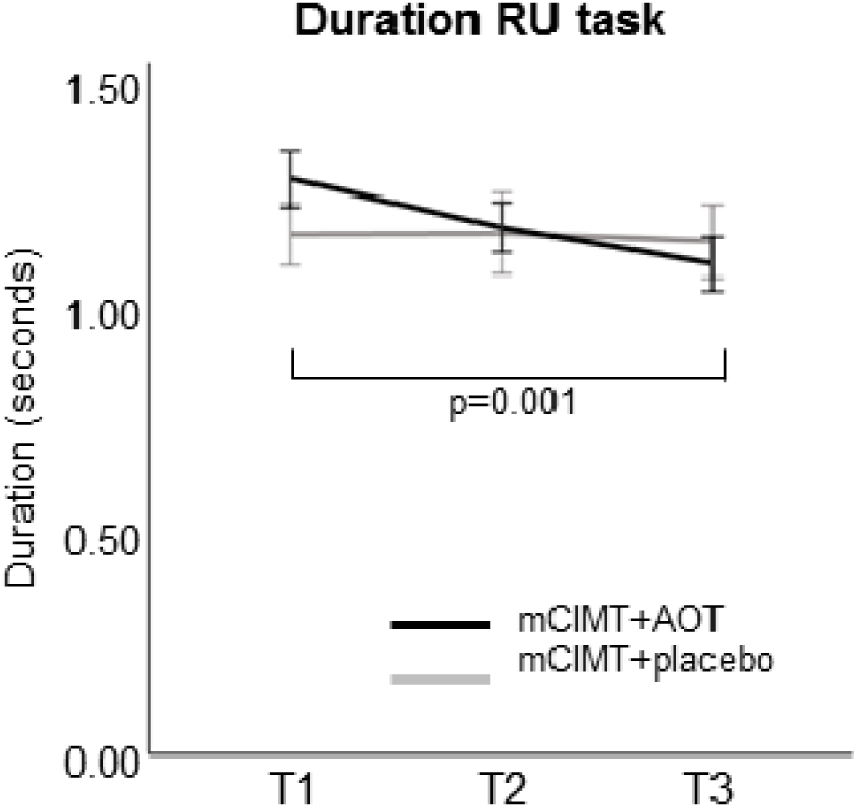
Movement duration during task reach upwards (RU). The mCIMT+AOT group improved more than the control group. Differences were significant at 6 months follow-up.

A summary of the findings based on the SPM1d analyses of the kinematic waveforms are reported in Table 3. We found time*group interactions for shoulder (all tasks), scapula (HTS), and wrist (HTS) kinematics, whilst no time*group interactions were found for the movement patterns of the trunk and elbow (p>0.05).

**Table 3.**
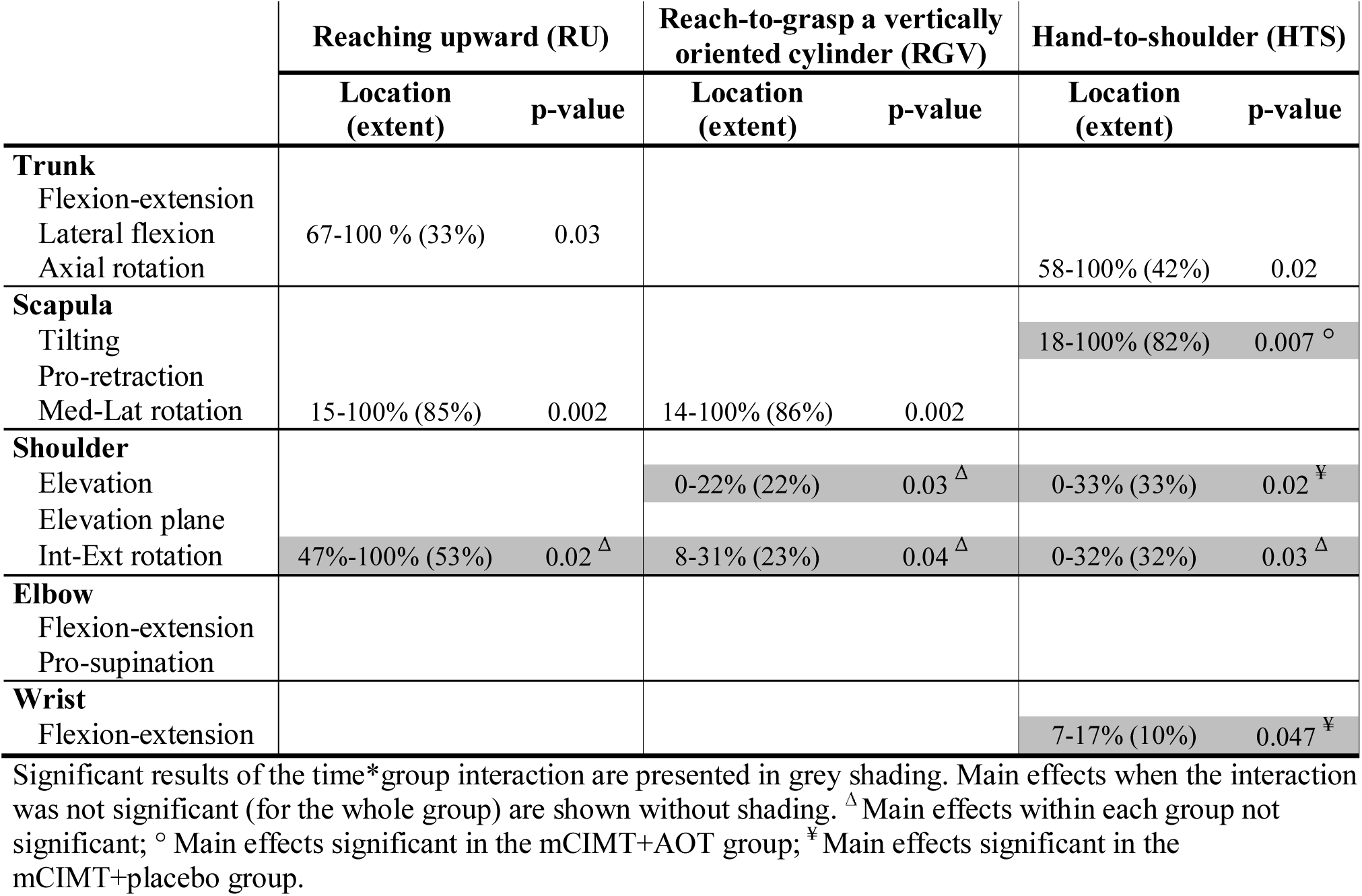
Statistical parametric mapping results of the effect of the intervention over time, for each joint angle, for the three tasks.

For the shoulder, further analysis showed no significant differences in any of the intervention groups for **shoulder rotation** (all tasks) and **shoulder elevation** (RGV). The analysis for **shoulder elevation** also showed a time*group interaction (HTS: p=0.02, 0-33% of the movement cycle), whereby the mCIMT+placebo group performed the task with less shoulder elevation over time (p=0.04, 11-24% of the movement cycle), whilst the mCIMT+AOT group did not show changes over time (p>0.05, Figure 3B). Post-hoc analyses in the mCIMT+placebo group indicated that the improvements in shoulder elevation occurred between T1-T3 (p=0.04, 8-24% of the movement cycle), but were not significant after Bonferroni correction (p>0.017).

**Figure 3.**
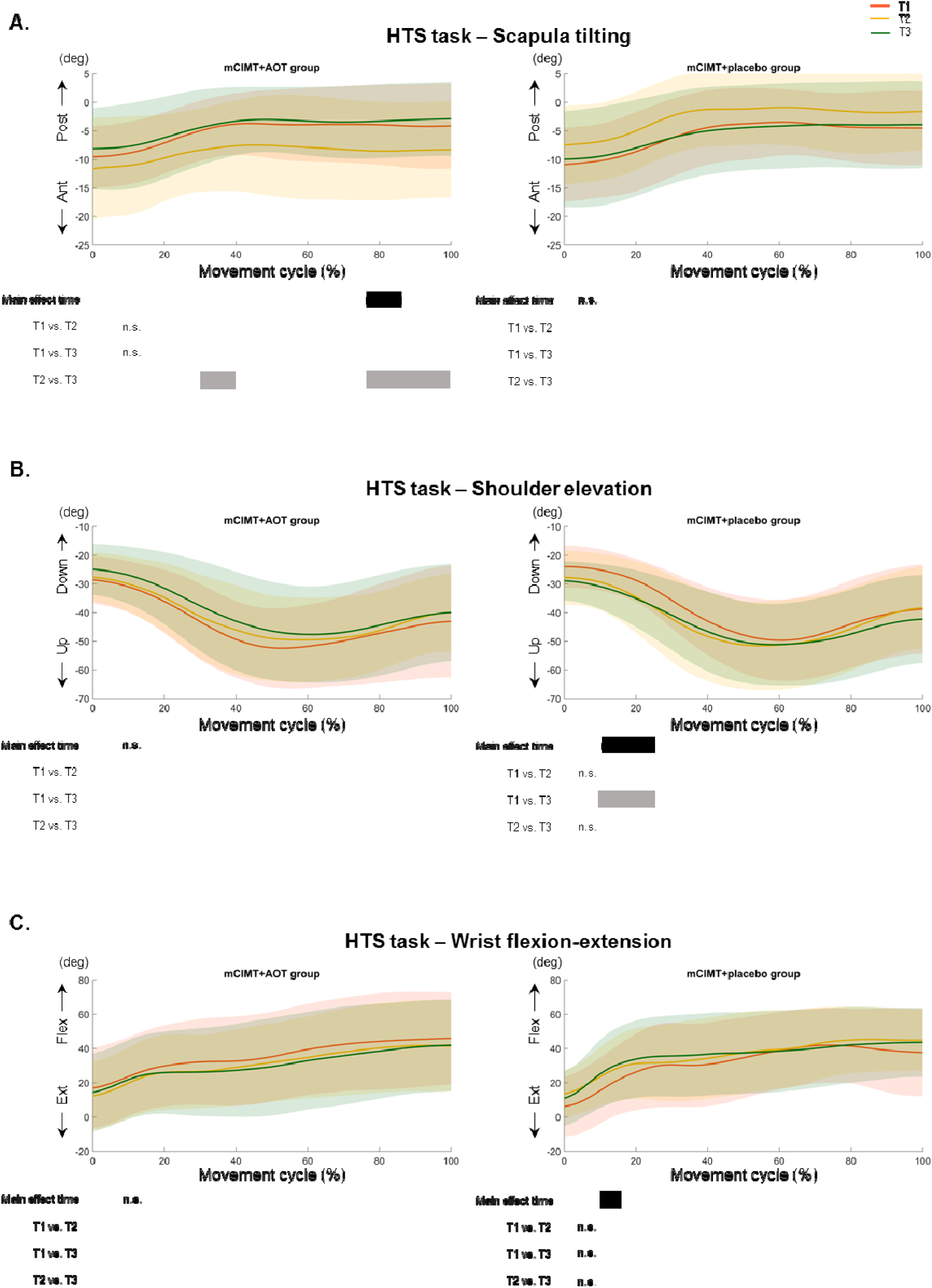
Time*group interactions for (A) scapula tilting, (B) shoulder elevation, and (C) wrist flexion-extension during the performance of the hand-to-shoulder task. Mean and standard deviation of the mCIMT+AOT group (left panel) and mCIMT+placebo group (right panel) is shown. Each subpanel displays where over the waveform the main effects (black bars) and the post-hoc analyses (grey) in each group were depicted.

We found a time*group interaction for **scapula tilting** (HTS: p=0.007, 18-100% of the movement cycle), showing changes over time in the mCIMT+AOT group (p<0.05, 77-85% of the movement cycle). The increased anterior tilting of the scapula after the intervention (i.e. increased pathological pattern) normalized again at T3 (Figure 3A).This change was significant only between T2-T3 (cluster 1: p=0.02, 31-41% of the movement cycle; cluster 2: p=0.01, 76-100% of the movement cycle). It is important to note that the interaction for this joint angle may have been biased by the increased anterior tilting in the mCIMT+AOT group whilst the mCIMT+placebo group showed decreased anterior tilting (Figure 3A). Supplementary Figures S1-S12 show the waveforms at different time points, for each group and for the total group, with additional plotting of normative data from typically developing children from Simon-Martinez et al (2018) ^26^.

Lastly, we also found a time*group interaction for **wrist flexion-extension** (HTS: p<0.05, 7-17% of the movement cycle). Subsequent analysis in each group indicated increased wrist flexion in the mCIMT+placebo group (p<0.05, 10-16% of the movement cycle). Although this was not significant in the post-hoc analyses (p>0.05), visual inspection indicated that this occurred between T1-T2 and was maintained at T3 (Figure 3C).

### Within-group analysis

Both groups improved over time in **trajectory straightness and peak velocity** (Figure 4). Trajectory straightness improved during HTS (p=0.04), and a trend was also found for RU and RGV (p=0.07) with large effect sizes (partial η^2^=0.15 for both tasks, Figure 4). Post-hoc analyses depicted that these changes occurred between T1-T3 and T2-T3, although they were not significant (p=0.07, p=0.09, respectively; Figure 4). Similar results were found for peak velocity, which significantly changed over time during RU (p=0.01) and RGV (p=0.008), also with large effect sizes (partial η^2^= 0.23-0.25; Figure 4). Post-hoc analyses for RU showed significant improvements between T1-T3 (p=0.02) and T2-T3 (p=0.03). Improvements during RGV were found immediately after the intervention (T1-T2,p=0.04) and maintained at follow-up (T1-T3, p=0.02). No improvements over time were found for **time-to-peak velocity** (all tasks, p>0.05; Figure 4).

**Figure 4.**
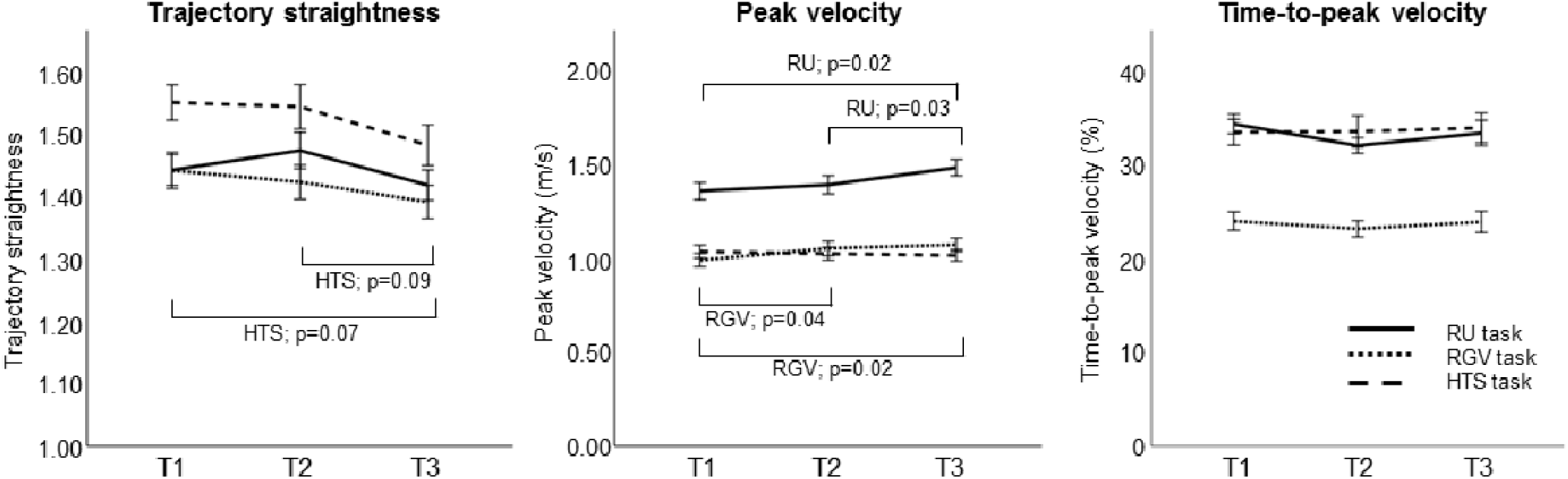
Change in spatiotemporal parameters over time for each task: reach upwards (RU), reach-to-grasp a vertically oriented cylinder (RGV), and hand-to-shoulder (HTS). Lines indicate mean and standard errors.

A summary of the results of the main effects are reported in Table 3. At the proximal level we found changes in **trunk** axial rotation for HTS (p=0.02, 58-100% of the movement cycle). Post-hoc analysis showed a trend toward increased external rotation (normalizing the movement pattern) after the intervention (T1-T2, p=0.03, 64-100% of the movement cycle), which was not maintained at follow up (T1-T3, p>0.05; T2-T3, p=0.01, 72-100% of the movement cycle) (Figure 5A). We also found improved lateral flexion during RU (p=0.03, 67-100% of the movement cycle), although post-hoc analyses did not survive Bonferroni correction (Figure 5B).

**Figure 5.**
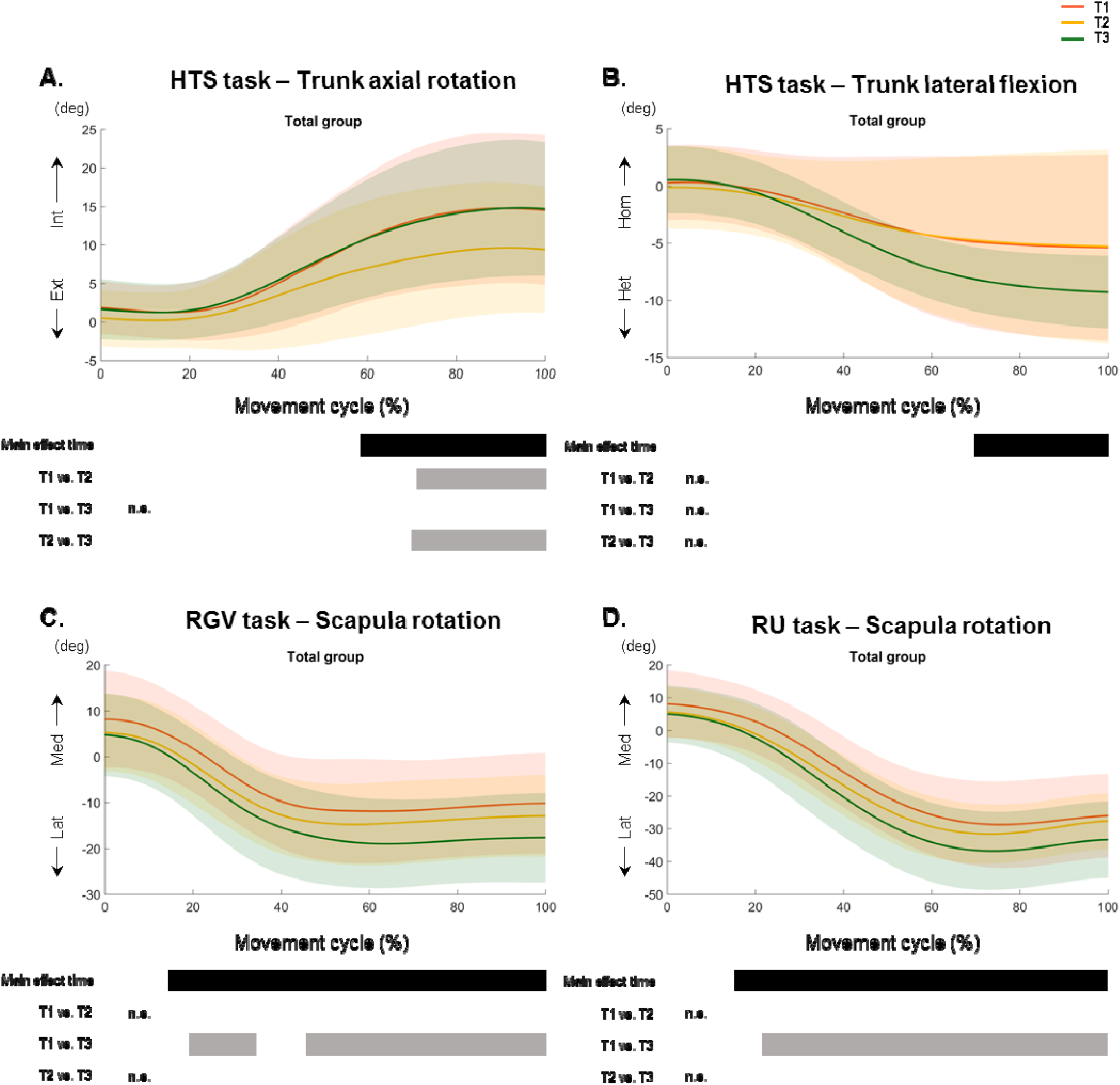
Change in movement pattern for (A) trunk axial rotation, (B) trunk lateral flexion, and (C-D) scapula rotation at T1, T2 and T3 for all participants. Mean and standard deviation for the total group is shown. Each subpanel displays where over the waveform the main effects (black bars) and the post-hoc analyses (grey) were depicted.

**Scapula medial-lateral rotation** improved for both reaching tasks (RU: p=0.002, 15-100% of the movement cycle; RGV: p=0.002, 14-100% of the movement cycle). Post-hoc analyses showed no differences immediately after the intervention (T1-T2, p>0.05), but the children used increased lateral rotation at follow-up for RU (T1-T3, p<0.001, 22-100% of the movement cycle; Figure 5D) and RGV (T1-T3, cluster 1: p=0.01, 19-36% of the movement cycle; cluster 2: p=0.003, 45-100% of the movement cycle; Figure 5C), resembling the usual movement pattern in typically developing children. There were no changes at the level of the **shoulder, elbow** or **forearm** (p>0.05).

## 3. Discussion

This is the first study investigating the effect of an intensive training approach combining mCIMT and AOT on UL motor control and movement patterns in a large cohort of children with uCP. The evaluation included a comprehensive UL kinematic analysis including spatiotemporal parameters and kinematic joint angles. Furthermore, the analysis with SPM1d permitted an investigation on changes in UL movement patterns over the entire waveform. Our results showed that the additional value of AOT was rather small, as measured with 3DMA. The mCIMT+AOT group performed RU significantly faster than the control group. As to UL movement patterns, we found between-groups differences mainly between T2-T3 with inconsistent results between groups. For the total group, we found an improved movement efficiency (peak velocity), a straighter trajectory, and improvements in trunk rotation and lateral flexion as well as in scapula rotation. These results highlight the importance of utilizing 3DMA to measure changes in proximal joints after an UL intervention.

### What is the added value of AOT to mCIMT on UL kinematics?

The combination of mCIMT+AOT resulted in a shorter movement time during a reaching task compared to those who did not receive additional AOT. The effect of the video-observation may have paced the rhythm of movement performance. The further decrease at T3 suggests that the added AOT training may have a long-lasting effect on movement duration during reaching. However, we did not collect information related to the child’s UL therapy between T2-T3 and this should be interpreted with caution. The only study investigating the effect of AOT on UL kinematics compared to placebo was performed in adult stroke survivors during a period of six weeks ^27^. In contrast to our study results, they reported improvements in both groups in average velocity and trajectory straightness after the intervention, without between-groups differences in outcomes.

Children who received mCIMT+AOT showed increased anterior scapula tilting after the intervention, which indicated an increased movement pathology ^25^. Given that the AOT videos only focused on the hand and the forearm, these increased proximal changes were unexpected and may have occurred at the expense of an improved grasp, as the children may have focused on improving their grasp function potentially leading to proximal compensations. However, grasp function is not assessed with our 3DMA protocol and future studies should include a kinematic evaluation of the grasp after an intervention, together with an evaluation of proximal joints. The abovementioned preliminary study ^27^ also pointed toward a limited immediate effect of AOT on UL kinematics in stroke survivors ^27^, which is in agreement with our results. Altogether, our results together with those reported by ^27^, suggest that AOT may have limited potential in improving the quality of the UL movement patterns in neurological populations.

### What are the effects of mCIMT (with or without AOT) on UL kinematics?

After mCIMT, with or without AOT, children generally performed the UL tasks with higher peak velocity, and a straighter trajectory, which may reflect improved motor control, resulting from the intensive skill-based practice in daily life activities. Peak velocity improved in both groups immediately after the intervention, and gains were maintained at follow-up, which is in agreement with previous studies in uCP after mCIMT ^28,29^. Children with uCP typically have a lower peak velocity compared to typically developing children ^19^, which has been related to muscle strength deficits ^22^. Although we cannot directly conclude this from current study results, the improvement in peak velocity may reflect gains in muscle force, which is crucial for daily life activities. Time-to-peak velocity, which represents the movement strategy, did not improve after the intervention. It is known that children with uCP usually do not show impairments in movement strategy ^19,30^, hence little improvement of this parameter was expected. Although it is possible that the tasks that we used were too simple to depict changes in this parameter. We also found gains in trajectory straightness, which was more pronounced at follow-up. Robert et al (2013) did report improvements in trajectory straightness immediately after a task-oriented training program of 5 weeks in children with uCP ^31^. This may suggest that children with uCP may need more time to improve trajectory straightness and transfer it to their daily life activities.

Whilst improvements in spatiotemporal parameters after mCIMT have been reported, the uniqueness of this study lays within the additional measurement of UL movement patterns with an evaluation at three time points and its analyses over the entire waveform. We found improvements in trunk and scapula movement patterns for the whole group. More specifically, we found a normalization of the trunk rotation during HTS after mCIMT. Additionally, scapula medial-lateral rotation also normalized during RU and RGV after the intervention (i.e. increased lateral rotation). These proximal improvements may highlight an improved neuromuscular control of the surrounding muscles ^32^. Adequate proximal control is crucial for an appropriate use of the distal joints ^33^. To the best of our knowledge, changes in joint kinematics have not yet been evaluated after an mCIMT intervention in children with uCP and our results point toward the value of a kinematic evaluation to identify changes in movement patterns after an intensive training.

In contrast to our expectations, we did not find distal improvements after our intervention at the level of the elbow and wrist. Whilst some studies have shown changes at the elbow in children with uCP ^34^ and in adult stroke ^35–37^, other studies evaluating UL movement quality using e.g. the Melbourne Assessment ^38^ have reported no or limited improvements after mCIMT ^39,40^. These studies highlight the difficulties in changing the distal pathological movement pattern during daily life activities and that a longer training period may be necessary. Nevertheless, we did find training-induced changes at the proximal joints, especially during HTS task, highlighting the responsiveness to change of some 3DMA-derived motor control features (i.e. duration, peak velocity, and trajectory straightness).

In summary, our study showed improvements in motor control after mCIMT, which is in line with the clearly shown benefits for the distal UL measured with traditional clinical scales ^41^. However, 3DMA showed its ability and uniqueness in capturing changes in proximal movement patterns. Mailleux et al (2017) previously put forward the importance and advantages of combining both clinical and kinematic UL evaluations, as each contributes in a specific and unique fashion to the identification of UL problems ^22^. Integrating both will surely provide a more comprehensive picture of the training-derived changes and will help clinicians target decomposed problems.

There are some limitations in this study. This study’s sample size was calculated to find improvements larger than the smallest detectable difference in bimanual performance (measured with the Assisting Hand Assessment) as a primary outcome measure ^42^. However, a total sample of 36 children may be insufficient to find improvements in distal UL movement patters after a two-week intervention. A second limitation of this study relates to the UL-3DMA protocol, as it did not measure grasp aperture and closure. Other 3DMA protocols measuring grasp and finger movements ^29,43^ might be able to capture improvements at these levels. Finally, for both the spatiotemporal characteristics and the joint movement patterns, we found large individual variability in response to the intervention. Previous studies have also shown large variation in clinical outcome measures after an mCIMT intervention ^44^, suggesting that there might be sub-groups of children who respond better. Although this has not yet been explored with a kinematic outcome, further analysis could also focus on identifying responders based on movement characteristics.

Future studies could also include bimanual tasks to evaluate improvements in bimanual coordination and the transfer of skills from a unimanual to a bimanual level, which has received very little attention in research ^45^. The use of SPM1d in this study has shown its ability to map treatment-dependent changes in uCP and could be included also in studies including kinematics during bimanual tasks. In addition to bimanual coordination, it would be interesting to include muscle activity measures (i.e. electromyography), which will allow for a more comprehensive understanding of the changes at the muscle activity level. Furthermore, the investigation of muscle synergies has been shown to offer insights into the complexity of motor control in uCP ^46,47^.

In conclusion, adding AOT to mCIMT mainly affects movement duration during reaching, whereas little benefit is observed on movement patterns. However, independent of AOT, mCIMT improved motor control parameters and proximal movement patterns, which are important to stabilize the UL to exert a coordinated, effective and smooth movement. This study adds to the available literature on intensive therapies in uCP and highlights the importance of including 3DMA in the UL evaluation to capture changes in motor control at proximal joints. Furthermore, we contributed to the identification of specific outcome parameters and tasks sensitive to change, advancing the clinical implementation of UL-3DMA.

## 4. Materials and Methods

This RCT protocol has been previously described in detail ^42^, and will be briefly summarized here. The study was conducted at KU Leuven and was approved by the Ethics Committee Research UZ / KU Leuven (S56513). All participants assented to participate, and their parents or caregivers signed the informed consent. The study is registered at www.clinicaltrials.gov (identifier NCT03256357).

### 4.1. Study population and randomization

Children with uCP aged 6-12 years were recruited via the CP reference centre of the University Hospitals Leuven. Children were included if they had a score between 4 (i.e. poor active assist) and 8 (i.e. spontaneous use) according to the House Functional Classification (HFC) scale ^48^, and sufficient cooperation to complete the test procedure. Exclusion criteria were UL surgery 2 years prior to testing or UL botulinum toxin A injections 6 months prior to testing. Between 2014 and 2017, we organized seven summer camps in which the children were prospectively enrolled. Participants were first stratified according to the HFC scale (4–5 vs. 6–7), age (6-9y vs. 10–12 y), and the type of corticospinal tract wiring pattern (contralateral, bilateral and ipsilateral as measured with transcranial magnetic stimulation). Next, participants were allocated to the experimental (mCIMT+AOT) or the control (mCIMT+placebo) group, by using a permuted block design of two.

## 4.2. Intervention

The intervention was delivered in a day camp model for 9 out of 11 consecutive days (6 hours/day, total of 54 hours of therapy), with no therapy during the weekend. To guarantee individual guidance, the therapist-child ratio was 1:1. A team of experienced paediatric physiotherapists led the camps, assisted by paediatric physiotherapy master students. During the camp hours, all children wore a tailor-made hand splint on the less impaired hand while performing unimanual exercises during individual therapy or group activities, and during the AOT or placebo condition.

The **individual therapy** (9h) was based on motor learning principles of shaping and repetitive practice focussing on: (1) active wrist and elbow extension, (2) forearm supination, (3) grip strength, and (4) fine motor tasks. These goals were embedded in functional tasks, which were tailored to meet the child’s therapeutic needs and adapted according to the child’s progress. The **group activities** (30h) consisted of painting, crafting, cooking, and outdoor games, specifically selected to stimulate the intensive use of the more-impaired hand. Children assigned to the mCIMT+AOT group received 15 **AOT sessions** (each lasting 1 hour), during which they watched video sequences showing unimanual goal-directed actions from the first-person perspective. All actions were adapted to the UL functional level of the child (see Additional files 1 and 2 of ^42^). The action was observed for 3 minutes and afterwards executed for 3 minutes, which was repeated twice per action, totalling six actions per session. The children in the mCIMT+placebo group played video games without biological motion (15h). Afterwards, the therapist explained the action and the child executed it for 3 minutes. To assure attention, a yes/no question related to each video activity was asked after the second execution of each activity (e.g. is the box taken from the top? Did you see the palm of the hand?). At the end of the intervention, the number of correct answers were summed up, ranging from 0 (all answers incorrect) to 45 (all answers correct).

### 4.3. Evaluation

The evaluation consisted of an UL-3DMA, based on a protocol specially developed for children with uCP ^21,22^. The protocol has shown to be reliable and valid, and full protocol details can be found elsewhere ^21,42^. Seventeen reflective markers were mounted on the trunk, acromion, upper arm, forearm and hand to record UL motion with 12-15 Vicon infrared cameras (Oxford Metrics, Oxford, UK) sampling at 100 Hz. Children sat in a custom-made chair that allowed standard sitting with foot and back support. We identified the anatomical landmarks of interest with static calibration trials following the International Society of Biomechanics guidelines ^49^. The movement protocol included three tasks: reaching upward (RU), reach-to-grasp vertical (RGV), and hand-to-shoulder (HTS), which are highly discriminative in children with uCP ^25^. Children were instructed to perform each task four times at self-selected speed, in two different trials, resulting in 8 movement repetitions per task. Children were evaluated right before (T1) and after (T2) the intervention, as well as at 6 months follow-up (T3). LM and CSM conducted the UL-3DMA evaluations.

### 4.4. Data processing

Offline data processing was performed using Vicon Nexus software (version 1.8.5, Oxford Metrics, Oxford, UK) and included a Woltring filtering routine with a predicted mean squared error of 10 mm^2 50^, gap filling, and selection of the movement cycles (start and endpoints). Only the middle two repetitions of each trial were time-normalized (0-100%) and used for the kinematic analysis. The open source software U.L.E.M.A v1.1.9 ^21,51,52^ was used to retain the three cycles with the lowest root mean square error for the computation of spatiotemporal parameters and joint kinematics. Spatiotemporal parameters were extracted from the central hand marker and included movement duration (seconds), peak velocity (m/s), time-to-peak velocity (% of the cycle), and trajectory straightness (unit less). A higher peak velocity indicates greater force or impulse during the movement. An earlier time-to-peak velocity points to the used movement strategy, dividing the movement in ‘before’ (time spent during the first visually triggered outward movement) and ‘after’ the peak velocity (the second half of the movements requires more precision to be successful) ^53^. Lastly, trajectory straightness indicates movement efficiency. Kinematic joint angles were calculated for the trunk (axial rotation, lateral flexion, and flexion-extension), scapula (medial-lateral rotation, tilting, and pro-retraction), shoulder (internal-external rotation, elevation plane, and elevation), elbow (flexion-extension and pro-supination), and wrist (flexion-extension).

### 4.5. Statistical analyses

Spatiotemporal parameters were checked with the Shapiro-Wilk test and their histograms were checked for symmetry. Descriptive statistics are reported as mean and standard errors of the mean. A repeated-measures Analysis of Variance (rmANOVA) was conducted to evaluate changes over time (T1, T2, and T3) and between groups (mCIMT+AOT vs. mCIMT+placebo). The time*group interaction was included in the model to explore potential different time trends between groups. In case of a significant interaction, time trends were investigated separately per group. Otherwise, time trends were further investigated in the whole group. The alpha-level for interaction and main effects for the spatiotemporal parameters was set at 0.05, with a post-hoc Tukey HSD correction. Effect sizes are reported for the comparison of the spatiotemporal parameters as partial η^2^ in the rmANOVA models (small 0.01; medium 0.06; large 0.14) ^54,55^ and as Cohen’d effect sizes in the pair-wise comparisons (small, 0.2–0.5; medium, 0.5–0.8; large>0.8) ^55^. The statistical analysis of the spatiotemporal data was performed in SPSS (version 25.0, SPSS Inc, Chicago, IL, USA).

The waveform analysis of joint angles was conducted using the Statistical Parametric Mapping (SPM) for 1 dimensional data toolbox (SPM1d, version 0.4 for Matlab, available for download at http://www.spm1d.org/Downloads.html) ^56^. SPM1d allows for hypothesis testing over the entire spectrum by considering the interdependency of the data points using random field theory and thus reduces the risk of type I errors. For every joint angle, the waveforms are compared using the conventional univariate statistic of an rmANOVA, outputting a statistical curve (F-curve). Next, random field theory is applied to estimate the critical threshold above which only 5% of equally smoothed random data is expected to cross (α<0.05). When clusters (i.e. differences between groups or over time) are identified, the location, extent, and a single p-value is extracted. Clusters smaller than 5% of the movement cycle are not reported due to little clinical relevance. For every joint angle, we first tested the time*group interaction to evaluate different time trends between groups. If the interaction was significant, time trends were investigated in each group. Otherwise, time trends were investigated for the whole group. For the SPM1d post-hoc comparisons, alpha-level for interaction and main effects was set at 0.05, and Bonferroni-corrected for post-hoc analysis (division by the number of comparisons (i.e. 0.05/3)).

## Data Availability

Data is available upon request to the corresponding author

## Data Availability

Data is available upon request to the corresponding author

## 5. Data availability

Data related to the manuscript is available upon request to corresponding author.

## 7. Acknowledgments

We would like to express our deepest gratitude to the children and families who participated in this study. We especially thank Catherine Huenaerts and Davide Monari for their assistance with data collection and update of ULEMA software, respectively. We are also grateful to the students of the master in rehabilitation sciences who participated in the summer camps and assisted in the measurements.

## 8. Author contributions

HF, KK, EO and KD conceptualized the study. CSM and LM ran the study and conducted the kinematic evaluations. CSM and LM curated the data, with the assistance of EJ. CSM conducted the statistical analysis of the data and wrote the paper. HF and KK supervised the study. All authors reviewed, edited and approved the manuscript.

## 9. Competing interests statement

The authors declare that there is no conflict of interest regarding the publication of this paper.

## 10. Funding statement

This work is funded by the Fund Scientific Research Flanders (FWO-project, grant G087213N) and by the Special Research Fund, KU Leuven (OT/14/127, project grant 3M140230).

## Notes

### Competing Interest Statement

The authors have declared no competing interest.

### Clinical Trial

NCT03256357

### Author Declarations

All relevant ethical guidelines have been followed and any necessary IRB and/or ethics committee approvals have been obtained.

Any clinical trials involved have been registered with an ICMJE-approved registry such as ClinicalTrials.gov and the trial ID is included in the manuscript.

## References

1. Sakzewski, L., Boyd, R. & Ziviani, J. Clinimetric properties of participation measures for 5- to 13-year-old children with cerebral palsy: A systematic review. Developmental Medicine and Child Neurology 49, 232–240 (2007).

2. Gordon, A. M., Schneider, J. A., Chinnan, A. & Charles, J. R. Efficacy of a hand-arm bimanual intensive therapy (HABIT) in children with hemiplegic cerebral palsy: A randomized control trialDevelopmental Medicine and Child Neurology 49, 830–838 (2007).

3. Hoare, B., Imms, C., Carey, L. & Wasiak, J. Constraint-induced movement therapy in the treatment of the upper limb in children with hemiplegic cerebral palsy: a Cochrane systematic review. Clinical Rehabilitation 21, 675–685 (2007).

4. Novak, I., Cusick, A. & Lannin, N. Occupational therapy home programs for cerebral palsy: double-blind, randomized, controlled trialPediatrics 124, e606–14 (2009).

5. Novak, I. et al. A systematic review of interventions for children with cerebral palsy: state of the evidence. Developmental medicine and child neurology 55, 885–910 (2013).

6. Sakzewski, L., Ziviani, J. & Boyd, R. Systematic Review and Meta-analysis of Therapeutic Management of Upper-Limb Dysfunction in Children With Congenital HemiplegiaPEDIATRICS 123, e1111–e1122 (2009).

7. Charles, J., Lavinder, G. & Gordon, A. M. Effects of constraint-induced therapy on hand function in children with hemiplegic cerebral palsy. Pediatric physical therapyl: the official publication of the Section on Pediatrics of the American Physical Therapy Association 13, 68–76 (2001).

8. Sakzewski, L. et al. Randomized trial of constraint-induced movement therapy and bimanual training on activity outcomes for children with congenital hemiplegia. Developmental medicine and child neurology 53, 313–20 (2011).

9. Klingels, K. et al. Randomized Trial of Modified Constraint-Induced Movement Therapy With and Without an Intensive Therapy Program in Children With Unilateral Cerebral Palsy. Neurorehabilitation and Neural Repair 27, 799–807 (2013).

10. Taub, E., Uswatte, G. & Pidikiti, R. Constraint-Induced Movement Therapy: a new family of techniques with broad application to physical rehabilitation--a clinical review. Journal of rehabilitation research and development 36, 237–51 (1999).

11. Steenbergen, B., Verrel, J. & Gordon, A. M. Motor planning in congenital hemiplegia. Disability and Rehabilitation 29, 13–23 (2007).

12. Mutsaarts, M., Steenbergen, B. & Bekkering, H. Anticipatory planning deficits and task context effects in hemiparetic cerebral palsyExperimental Brain Research 172, 151–162 (2006).

13. Rizzolatti, G. & Craighero, L. THE MIRROR-NEURON SYSTEM. Annual Review of Neuroscience 27, 169–192 (2004).

14. Kim, J., Kim, J. & Ko, E. The effect of the action observation physical training on the upper extremity function in children with cerebral palsyJournal of Exercise Rehabilitation 10, 176–183 (2014).

15. Sgandurra, G. et al. Randomized Trial of Observation and Execution of Upper Extremity Actions Versus Action Alone in Children With Unilateral Cerebral Palsy. Neurorehabilitation and Neural Repair 27, 808–815 (2013).

16. Kirkpatrick, E., Pearse, J., James, P. & Basu, A. Effect of parent-delivered action observation therapy on upper limb function in unilateral cerebral palsy: a randomized controlled trialDevelopmental Medicine and Child Neurology 58, 1049–1056 (2016).

17. Buccino, G. et al. Action Observation Treatment Improves Upper Limb Motor Functions in Children with Cerebral Palsy: A Combined Clinical and Brain Imaging Study. Neural Plasticity 2018, 1–11 (2018).

18. Buccino, G. et al. Improving upper limb motor functions through action observation treatment: A pilot study in children with cerebral palsy. Developmental Medicine and Child Neurology 54, 822–828 (2012).

19. Jaspers, E. et al. Three-dimensional upper limb movement characteristics in children with hemiplegic cerebral palsy and typically developing children. Research in Developmental Disabilities 32, 2283–2294 (2011).

20. Jaspers, E et al. The reliability of upper limb kinematics in children with hemiplegic cerebral palsy Gait and Posture 33, 568–575 (2011).

21. Jaspers, E. et al. Upper limb kinematics: Development and reliability of a clinical protocol for children. Gait & Posture 33, 279–285 (2011).

22. Mailleux, L et al. Clinical assessment and three-dimensional movement analysis: An integrated approach for upper limb evaluation in children with unilateral cerebral palsy PLOS ONE 12, e0180196 (2017).

23. Fitoussi, F. et al. Upper limb motion analysis in children with hemiplegic cerebral palsy: Proximal kinematic changes after distal botulinum toxin or surgical treatments. Journal of Children’s Orthopaedics 5, 363–370 (2011).

24. Mackey, A. H., Miller, F., Walt, S. E., Waugh, M. C. & Stott, N. S. Use of three-dimensional kinematic analysis following upper limb botulinum toxin a for children with hemiplegia European Journal of Neurology 15, 1191–1198 (2008).

25. Simon-Martinez, C. et al. Negative Influence of Motor Impairments on Upper Limb Movement Patterns in Children with Unilateral Cerebral Palsy. A Statistical Parametric Mapping Study. Frontiers in Human Neuroscience 11, 482 (2017).

26. Simon-Martinez, C et al. Age-related changes in upper limb motion during typical development PLOS ONE 13, e0198524 (2018).

27. Kim, E. & Kim, K. Effects of purposeful action observation on kinematic patterns of upper extremity in individuals with hemiplegia. Journal of Physical Therapy Science 27, 1809–1811 (2015).

28. Chen, H. C. et al. Younger children with cerebral palsy respond better than older ones to therapist-based constraint-induced therapy at home on functional outcomes and motor control. Physical and Occupational Therapy in Pediatrics 36, 171–185 (2016).

29. Chen, H. C. et al. Improvement of upper extremity motor control and function after home-based constraint induced therapy in children with unilateral cerebral palsy: Immediate and long-term effects. Archives of Physical Medicine and Rehabilitation 95, 1423–1432 (2014).

30. Mackey, A. H., Walt, S. E. & Stott, N. S. Deficits in upper-limb task performance in children with hemiplegic cerebral palsy as defined by 3-dimensional kinematics. Archives of Physical Medicine and Rehabilitation 87, 207–215 (2006).

31. Robert, M. T., Guberek, R., Sveistrup, H. & Levin, M. F. Motor learning in children with hemiplegic cerebral palsy and the role of sensation in short-term motor training of goal-directed reaching Developmental Medicine and Child Neurology 55, 1121–1128 (2013).

32. De Baets, L., Jaspers, E., Janssens, L. & Van Deun, S. Characteristics of Neuromuscular Control of the Scapula after Stroke: A First Exploration. Frontiers in Human Neuroscience 8, 933 (2014).

33. Case-Smith, J., Fisher, A. G. & Bauer, D. An analysis of the relationship between proximal and distal motor control. The American journal of occupational therapy.l: official publication of the American Occupational Therapy Association (1989). doi:10.5014/ajot.43.10.657

34. Cimolin, V. et al. Constraint-induced movement therapy for children with hemiplegia after traumatic brain injury: a quantitative study. The Journal of head trauma rehabilitation 27, 177–187 (2012).

35. Wu, C.-Y. et al. Pilot trial of distributed constraint-induced therapy with trunk restraint to improve poststroke reach to grasp and trunk kinematics. Neurorehabilitation and neural repair 26, 247–255 (2012).

36. Woodbury, M. L. et al. Effects of trunk restraint combined with intensive task practice on poststroke upper extremity reach and function: a pilot study. Neurorehabilitation and neural repair 23, 78–91 (2009).

37. Michaelsen, S. M., Dannenbaum, R. & Levin, M. F. Task-specific training with trunk restraint on arm recovery in stroke: randomized control trial Stroke 37, 186–192 (2006).

38. Randall, M., Imms, C., Carey, L. M. & Pallant, J. F. Rasch analysis of the Melbourne assessment of unilateral upper limb function. Developmental Medicine and Child Neurology 56, 665–672 (2014).

39. Islam, M. et al. Is outcome of constraint-induced movement therapy in unilateral cerebral palsy dependent on corticomotor projection pattern and brain lesion characteristics? Developmental Medicine and Child Neurology 56, 252–258 (2014).

40. Aarts, P. B., Jongerius, P. H., Geerdink, Y. A., van Limbeek, J. & Geurts, A. C. Effectiveness of Modified Constraint-Induced Movement Therapy in Children With Unilateral Spastic Cerebral Palsy: A Randomized Controlled Trial Neurorehabilitation and Neural Repair 24, 509–518 (2010).

41. Sakzewski, L., Gordon, A. & Eliasson, A.-C. The State of the Evidence for Intensive Upper Limb Therapy Approaches for Children With Unilateral Cerebral Palsy. Journal of Child Neurology 29, 1077–1090 (2014).

42. Simon-Martinez, C et al. Combining constraint-induced movement therapy and action-observation training in children with unilateral cerebral palsy: a randomized controlled trial BMC Pediatrics 18, 250 (2018).

43. Kuhtz-Buschbeck, J. P., Stolze, H., Jöhnk, K., Boczek-Funcke, A. & Illert, M. Development of prehension movements in children: a kinematic study. Experimental Brain Research 122, 424–432 (1998).

44. Hoare, B. & Greaves, S. Unimanual versus bimanual therapy in children with unilateral cerebral palsy: Same, same, but different. Journal of pediatric rehabilitation medicine 10, 47–59 (2017).

45. Hung, Y.-C., Casertano, L., Hillman, A. & Gordon, A. M. The effect of intensive bimanual training on coordination of the hands in children with congenital hemiplegia Research in Developmental Disabilities 32, 2724–2731 (2011).

46. Tang, L. et al. Assessment of Upper Limb Motor Dysfunction for Children with Cerebral Palsy Based on Muscle Synergy Analysis. Frontiers in Human Neuroscience 11, 130 (2017).

47. Goudriaan, M. et al. Non-neural Muscle Weakness Has Limited Influence on Complexity of Motor Control during Gait. Frontiers in Human Neuroscience 12, 5 (2018).

48. House, J. H., Gwathmey, F. W. & Fidler, M. O. A dynamic approach to the thumb-in palm deformity in cerebral palsy J Bone Joint Surg Am 63, 216–225 (1981).

49. Wu, G. et al. ISB recommendation on definitions of joint coordinate systems of various joints for the reporting of human joint motion—Part II: shoulder, elbow, wrist and hand. Journal of Biomechanics 38, 981–992 (2005).

50. Woltring, H. J. Smoothing and differentiation techniques applied to 3-D data. Three-dimensional analysis of human movement. (Human Kinetics, 1995).

51. Jaspers, E et al. The Arm Profile Score: A new summary index to assess upper limb movement pathology Gait & Posture 34, 227–233 (2011).

52. Upper Limb Evaluation in Motion Analysis (U.L.E.M.A.). Available at: https://github.com/u0078867/ulema-ul-analyzer. (Accessed: 6th December 2017)

53. Alt Murphy, M. & Häger, C. K. Kinematic analysis of the upper extremity after stroke – how far have we reached and what have we grasped? Physical Therapy Reviews 20, 137–155 (2015).

54. Gravetter, F. & Wallnau, L. Statistics for the behavioral sciences. (Wadsworth, 2004).

55. Cohen, J. Statistical power analysis for the behavioral sciences. Hillsdale, NJ (Elsevier Science, 1988).

56. Pataky, T. C. Generalized n-dimensional biomechanical field analysis using statistical parametric mapping. Journal of Biomechanics 43, 1976–1982 (2010).

